# Development of a RAG-based Expert LLM for Clinical Support in Radiation Oncology

**DOI:** 10.1101/2025.09.16.25335813

**Authors:** Tingjun Liu, Xucheng Wang, Matthew Inkman, Julian C. Hong, Michael R. Waters, Jin Zhang

## Abstract

The ability of pre-trained large language models (LLMs) to rapidly master novel natural language processing tasks holds transformative potential. However, pre-trained LLMs often struggle to achieve high performance in specialized domains such as oncology and have the tendency to deliver incorrect information confidently (“hallucinate”), limiting their utility in such contexts. Retrieval-augmented generation (RAG) addresses this limitation by dynamically incorporating authoritative, domain-specific knowledge directly into the LLM’s inference process. This approach significantly enhances LLM performance without the typical requirement for extensive fine-tuning or retraining.

In this study, we demonstrate the exceptional performance of a minimalist RAG pipeline (without additional model fine-tuning) on radiation oncology board-style examinations. Leveraging a meticulously curated knowledge base sourced from Gunderson & Tepper’s *Clinical Radiation Oncology, Fifth Edition* and NCCN guidelines, our model substantially surpassed the performance of contemporary OpenAI models, achieving an outstanding accuracy of 91.5% on the 2021 American College of Radiology (ACR) TXIT examination. This result markedly exceeds the performance benchmarks set by previous LLM-based approaches in this field, which attained a maximum accuracy of 74%.

Crucially, our model exhibited robust self-awareness regarding its knowledge boundaries, overcoming a glaring weakness of pre-trained LLMs; questions answered incorrectly were reliably flagged with low confidence scores (mean 4.12/10 vs. 7.36/10 for correct answers), highlighting areas inadequately represented within the RAG knowledge base. This precise uncertainty estimation underscores RAG’s unique strength in enhancing not just accuracy, but also the reliability and interpretability of model outputs.

We demonstrate that integrating domain-specific knowledge via RAG significantly enhances large language model performance in radiation oncology, enabling reliable confidence scoring previously unattainable with pretrained LLMs. This scalable approach may be well-suited for clinical decision support and medical education. Future efforts will incorporate clinical guidelines and select primary literature to broaden applicability.

## 1 Introduction

Pre-trained large language models (LLMs) such as GPT-3 and GPT-4 have demonstrated remarkable potential in clinical medicine, generating human-like text and aiding in diverse tasks. Early studies showed that LLMs could pass specialist board examinations, assist in patient communication, and even draft scientific content^1, 2^. These capabilities foreshadow a disruptive impact on clinical practice and medical education. However, alongside their promise, general-purpose LLMs exhibit important limitations in healthcare settings. Chief among these is the tendency to produce hallucinations, i.e., plausible-sounding but false or unsubstantiated information^3, 4^. Without proper safeguards, an LLM might confidently generate incorrect clinical advice, thereby undermining trust and potentially endangering patients.

The limitations of current LLMs are especially pronounced in medical applications. General models are trained on broad internet text and may lack coverage of specialized, up-to-date medical knowledge^3^. In clinical usage, this can lead to grave issues: LLMs have been observed to misattribute symptoms to diseases or suggest improper diagnostic plans if critical contextual details are missing. They might also present outdated guidelines or inaccurate statistical claims, reflecting gaps or cutoffs in their training data^4^. This unreliability is problematic in any medical domain, but it is particularly acute in oncology, where the knowledge base evolves rapidly and errors in therapeutic recommendations can have life-threatening consequences. Indeed, a recent review of LLM applications in oncology noted significant error rates and instances of obsolete information in model-generated answers^5^. These observations underscore that off-the-shelf LLMs, though powerful, often fall short of the rigor and currency required for clinical decision support in oncology.

Within oncology, the subfield of radiation oncology presents unique challenges that further limit general LLM performance. Radiation oncology involves complex treatment planning, dose calculation, and adherence to technical protocols that demand a high level of precision and specialized knowledge. For example, designing a safe radiotherapy plan requires familiarity with detailed dose constraints, imaging modalities, and quality assurance measures rarely found in general text corpora. Small errors or omissions in this domain can directly affect patient safety. Not surprisingly, evaluations have shown that state-of-the-art general LLMs do not yet achieve expert-level competency on radiation oncology tasks without adaptation. Wang et al. (2025) found that base GPT-3.5 and GPT-4 models struggled on a standard medical physics exam, scoring well below the passing threshold for human experts^6^. This performance gap reflects the broader issue that general LLMs lack sufficient embedded knowledge of niche domains like radiotherapy physics and clinical oncology. They also cannot reliably update their knowledge base post-training, making it difficult to account for new evidence or guideline changes in oncology care. Taken together, these challenges highlight the need for strategies to bolster the domain-specific expertise and factual accuracy of LLMs in radiation oncology.

Retrieval-Augmented Generation (RAG) has emerged as a promising solution to address these limitations^3, 7^. RAG is a framework that integrates an external knowledge retrieval step into the LLM’s generation process. Instead of relying solely on its fixed internal parameters, a RAG-based model actively pulls in relevant information from a curated database or knowledge repository at query time. By providing the model with domain-specific context (e.g., excerpts from oncology guidelines or textbooks) before it produces an answer, RAG can ground the LLM’s responses in factual, up-to-date information. Prior work has shown that this approach significantly improves output accuracy and reduces hallucinations in knowledge-intensive tasks^8^. Essentially, RAG endows the LLM with a form of open-book exam capability: the model can consult trusted references on-the-fly, which is crucial in a field like oncology where veracity and currency of information are paramount. RAG also offers a practical advantage in that it does not require retraining the large model; instead, the domain knowledge is injected at inference time, allowing rapid updates and domain adaptation using new data^8^. This makes it an attractive strategy for clinical applications, where the knowledge base (e.g., clinical trial results, practice guidelines) is continually evolving.

In light of these considerations, we are motivated to develop an expert LLM for radiation oncology clinical support using a RAG-based approach. By integrating authoritative radiation oncology sources into the model’s retrieval corpus, we aim to create a system that combines the generative fluency of LLMs with the deep specialty knowledge of oncology experts. Key sources of truth, such as national clinical practice guidelines, radiation dose reference manuals, and peer-reviewed research articles, are leveraged to inform the model’s answers; for instance, prior researchers have compiled radiotherapy textbooks and professional society reports into a retrievable database to bolster a model’s medical physics knowledge^9^. The resulting RAG-enhanced LLM is designed to provide contextually relevant and factually grounded responses to questions in radiation oncology. Our goal is to achieve high accuracy and reliability comparable to human domain experts, while maintaining the efficiency and versatility of a language model. In the following, we present the framework and evaluation of this expert RAG-based LLM, which seeks to advance clinical decision support in radiation oncology by overcoming the limitations of general-purpose models.

## 2 Related Work

### LLMs in Medicine

The advent of transformer-based LLMs has catalyzed extensive research into their medical applications. Large models like GPT-3 and GPT-4 have demonstrated impressive performance on medical question-answering benchmarks, sparking interest in their potential for clinical decision support. For example, Kung et al. (2023) showed that an early version of ChatGPT (GPT-3.5) could perform at or near the passing threshold (i.e., 60% accuracy) on all three United States Medical Licensing Exam (USMLE) step exams without any domain-specific training^10^. GPT-4 has further pushed the envelope, reportedly achieving around 85-86% on USMLE-style questions, a level approaching expert physician performance^11^. Beyond general models, specialized medical LLMs have been developed to leverage domain data. Google’s Med-PaLM was the first LLM to exceed the USMLE passing score, and its successor Med-PaLM 2 combined medical fine-tuning with enhanced reasoning and retrieval techniques to reach 86.5% accuracy on a USMLE QA dataset^12^. Notably, in head-to-head evaluations, clinicians often preferred Med-PaLM 2’s answers to those written by human physicians^12^, highlighting the model’s potential to deliver high-quality clinical responses. Other initiatives like BioGPT^13^ and BioMedLM^14^ have trained language models on biomedical literature, yielding steady improvements on domain-specific benchmarks. These eNorts collectively demonstrate that while vanilla LLMs possess considerable medical knowledge, further adaptation – via fine-tuning or retrieval – greatly boosts their accuracy and usefulness in the medical domain.

### LLMs in Oncology

Given the success of LLMs in general medicine, researchers have explored their application in oncology, a domain with complex decision pathways and rapidly evolving knowledge. A recent systematic review by Carl et al. (2024) examined 34 studies of LLM use in clinical oncology^15^. The review found that LLMs (primarily ChatGPT and related models) have been tested on tasks ranging from answering oncology board-style questions and summarizing clinic notes to generating patient information handouts. Performance across these studies varied widely. In certain settings, LLMs demonstrated strong capabilities; for instance, providing sensible treatment options for common cancers or translating medical jargon for patients. However, heterogeneity in evaluation methods and prompt design led to a broad range of outcomes. Some studies reported near-expert accuracy, whereas others found significant lapses in the models’ oncologic knowledge or reasoning. Common challenges included the models’ tendency to offer outdated or incorrect information if the queries fell outside of well-represented training data. Indeed, despite overall promising results, Carl et al. noted that LLM responses in oncology often contained errors and occasionally reflected obsolete guidelines. This underlines a key issue: without explicit grounding in up-to-date cancer literature and guidelines, a general LLM may falter on specialist queries. As such, the oncology community has called for more robust, standardized benchmarks to fairly assess LLM performance and for techniques to improve the reliability of AI-generated answers in this domain.

### Radiation Oncology Q&A and Decision Support

Within oncology, radiation oncology has served as a challenging testbed for LLMs given its technical complexity. Thaker et al. (2024) conducted one of the first evaluations of LLMs in this field, assessing the performance of various models on the 2021 American College of Radiology (ACR) Radiation Oncology In-Training Examination (TXIT); they found that the best performing model, GPT-4-turbo, answered 74% of questions correctly^16^. Yalamanchili et al. (2024) prompted ChatGPT-3.5 with 115 radiation oncology patient-care questions curated from professional society websites^17^. The answers were compared against expert-written responses, and the results were cautiously optimistic. According to blinded radiation oncologists’ ratings, the LLM’s answers were on par or superior in 94% of cases for factual accuracy and in over 75% of cases for completeness. In 91% of the questions, the AI’s responses were more concise than the reference answers. These figures suggest that even a general LLM can capture a substantial portion of radiation oncology knowledge. However, the study also identified important concerns. In two instances, the model’s answer was flagged as potentially harmful, reflecting either a dangerous recommendation or a serious factual error. Moreover, the grade level of written responses from ChatGPT was significantly higher than that of the expert answers (mean grade level 13.6 vs 10.6), which could impair patient understanding. Thus, while LLMs show promise for answering radiotherapy questions, they require refinement to ensure patient safety and appropriate communication. Other works have assessed LLMs on more specialized radiation oncology tasks. Holmes et al. (2023) examined multiple foundation models on radiation oncology in-training exam questions and found notable variability in performance across models^18^. Crucially, they emphasized that advanced radiotherapy techniques (IMRT, proton therapy) pose a high bar for AI, given the need for precise numerical calculations and nuanced judgment. These domain-specific evaluations reinforce the notion that additional domain adaptation is needed for LLMs to function reliably as radiation oncology assistants.

### Retrieval-Augmented Models in Healthcare

To mitigate LLM knowledge gaps and hallucinations, researchers are increasingly turning to retrieval-based augmentation in the medical domain. The RAG paradigm, initially popularized in open-domain question answering, has been adapted for healthcare applications to inject current medical knowledge into model outputs. Grandinetti and McBeth (2023) introduced an Adaptable Retrieval-Based Chain-of-Thought (ARCoT) framework for medical physics, a subdiscipline of radiation oncology, as a notable example^9^. ARCoT combines a GPT-4 class LLM with a custom retrieval pipeline that draws on a library of vetted medical physics documents (e.g., AAPM task group reports and IAEA textbooks) to answer complex exam questions. With this approach, the authors reported that their augmented model outperformed the base LLMs by a large margin, even exceeding the average human physicist’s score on a challenging radiotherapy physics exam. Specifically, integrating retrieval and reasoning led to performance improvements up to 68%, with the hybrid model scoring about 90% on the test – well above what the unaugmented GPT-4 achieved. Importantly, the RAG-based system also produced far fewer physics mistakes, indicating a reduction in hallucinated answers and an increase in domain-specific accuracy. Similarly, other teams have shown that augmenting LLMs with clinical databases or guidelines can enhance their utility. Liu et al. (2023) incorporated oncology literature via a vector database to aid an LLM in generating radiotherapy treatment plans, finding that retrieval of relevant trial data improved the quality of the AI’s recommendations^3^. Such findings echo the consensus that access to curated knowledge is key for safe and effective AI in medicine. Nevertheless, challenges remain in optimizing RAG for healthcare, including managing the limited context window of LLMs and ensuring the retrieval module pinpoints the most relevant facts quickly. Ongoing research is exploring solutions like re-ranking algorithms to filter retrieved documents and refined prompting strategies (e.g., step-by-step reasoning prompts) to better utilize the fetched information. In summary, the integration of retrieval-based methods with LLMs is a promising approach, and our work builds upon this paradigm. By leveraging RAG with trusted radiation oncology sources, we aim to create an expert LLM system that aligns with clinical knowledge standards and supports oncology practitioners with reliable, evidence-based information.

## 3 Methodology

### 3.1 Overview of the RAG Pipeline

To systematically answer board-style radiation oncology questions using reliable, up-to-date evidence, we developed a multi-agent retrieval-augmented generation (RAG) pipeline, which is shown in **Figure 1**. Our pipeline is designed to leverage the strengths of large language models (LLMs) in language understanding and synthesis while ensuring that generated answers are grounded in authoritative reference materials and transparent to human reviewers.

**Figure 1:**
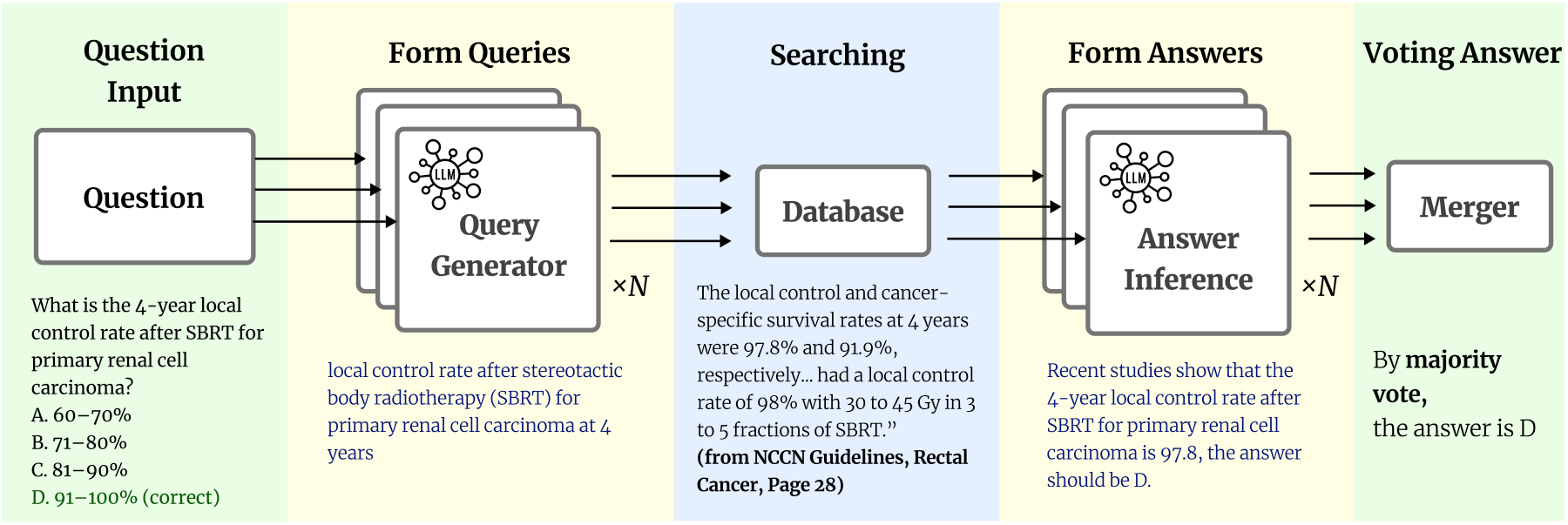
A schematic overview of our RAG-based workflow for answering radiation oncology questions. The system first generates multiple queries from the input question, retrieves relevant contexts from the reference database, and then uses answer inference and majority voting to produce the final answer.

#### Pipeline Workflow

Given a user query, such as “What is the 4-year local control rate after SBRT for primary renal cell carcinoma?”, the workflow consists of four steps:

1. The pipeline receives a multiple-choice or open-ended clinical question, automatically extracts key entities (disease, treatment, numbers, and time frame), and reformulates them into focused retrieval queries suitable for literature or guideline search. This modular design ensures that the subsequent search targets the most relevant information.
2. Each agent (*n*=5) generates a semantically rich retrieval query tailored for vector search in a curated medical database. For the example above, the generated query would be: “local control rate after stereotactic body radiotherapy (SBRT) for primary renal cell carcinoma at 4 years”.
3. The system retrieves evidence passages (e.g., from guidelines or recent publications), and each agent independently infers a structured answer based on the retrieved text. This answer includes both a natural-language explanation and, for multiple-choice settings, an explicit option selection (e.g., “D: 91–100%”). The use of LLMs for both retrieval and inference allows for nuanced interpretation and flexible reasoning.
4. All agent answers are aggregated using a majority vote. This ensemble strategy mitigates individual agent bias or errors, and provides a transparent audit trail for answer provenance. The final answer, along with supporting evidence, is then returned to the user.

#### Design Motivation

This multi-agent RAG architecture is motivated by the need for (a) accurate and up-to-date answers rooted in clinical best evidence, (b) explainability and traceability of LLM responses, and (c) robustness against individual retrieval or inference errors. By decomposing the process into discrete, auditable steps, our system enables users to receive high-confidence answers and review the supporting chain of evidence at each stage. The voting mechanism further increases reliability, especially in high-stakes medical settings where single-model hallucination or misinterpretation could have significant consequences.

### 3.2 Knowledge Base Construction

To ensure that the RAG pipeline can provide accurate, up-to-date, and clinically meaningful answers, we curated a comprehensive domain-specific knowledge base integrating several authoritative sources in radiation oncology (**Figure 2**). Specifically, the knowledge base consists of: Gunderson & Tepper’s *Clinical Radiation Oncology, Fifth Edition*^19^, the latest NCCN Guidelines relevant to radiation oncology^20^, and a supplementary set of 1,000 PubMed articles randomly selected using the query “Radiation Oncology”^21^. For each source, we performed text extraction from the original PDFs, followed by segmentation of the extracted text into contiguous chunks of 512 tokens. This chunk size was selected to balance semantic coherence and retrieval efficiency. To ensure provenance and transparency, each chunk was associated with rich metadata, including source type, document title, section heading, and page number, to allow for precise citation and traceability of any supporting evidence. All processed text chunks and their associated metadata were stored in a persistent ChromaDB vector database, forming the foundation for downstream retrieval and answer generation. This structured knowledge base enables the pipeline to perform fine-grained, context-aware retrieval from multiple evidence sources, and ensures that every answer generated can be traced back to an authoritative reference.

**Figure 2:**
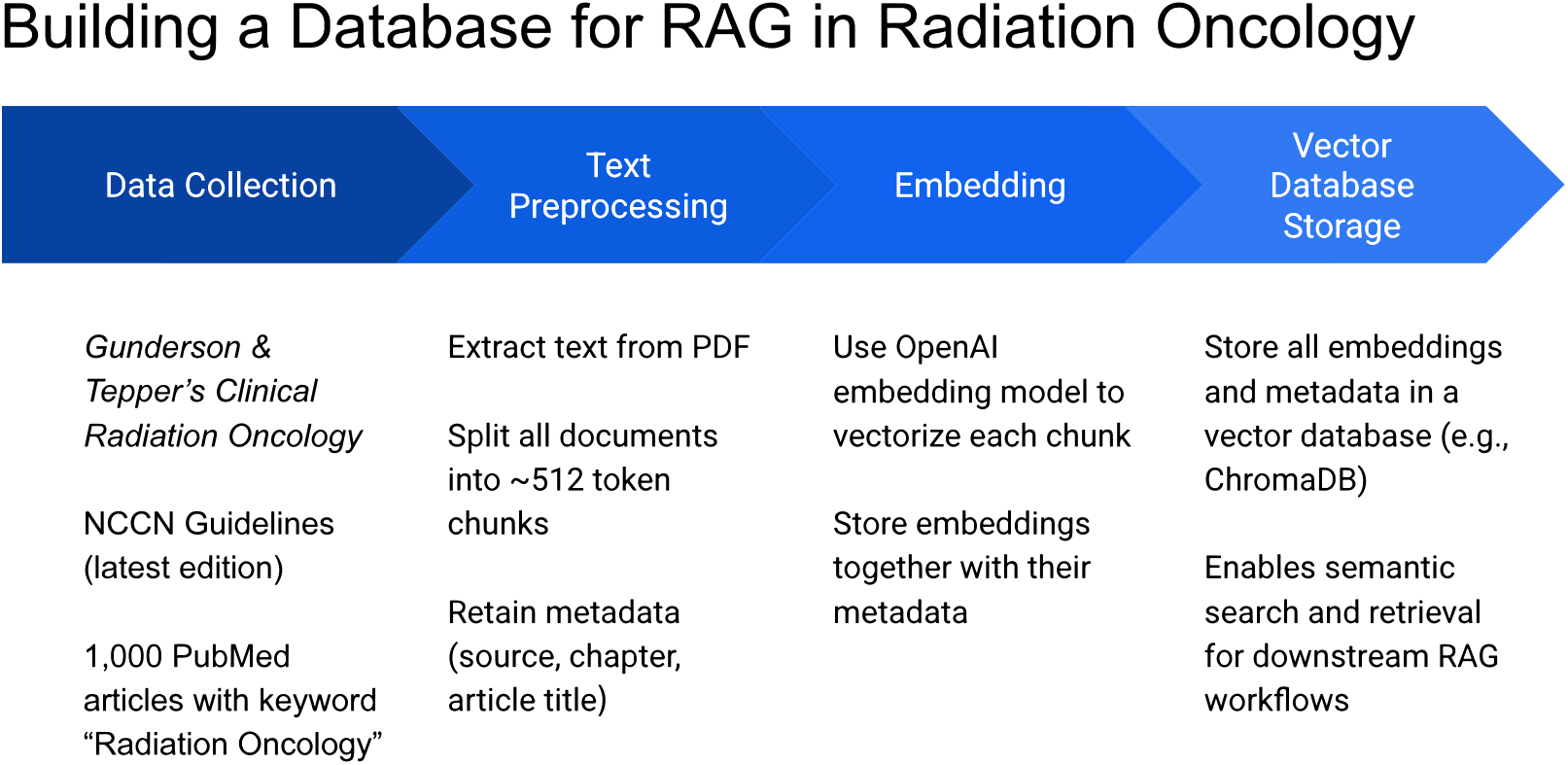
Overview of the domain-specific knowledge base construction pipeline. Authoritative sources are collected, preprocessed into semantically coherent text chunks with metadata, embedded into vector representations, and stored in a persistent vector database (ChromaDB) to enable accurate, context-aware retrieval and citation in downstream RAG workflows.

### 3.3 Retrieval Component

To ensure accurate and contextually relevant evidence retrieval, we utilized a two-step retrieval strategy based on dense vector similarity. Each input query, formulated as described in Section 3.1, is first embedded using OpenAI’s text-embedding-3-large model, a state-of-the-art language embedding model optimized for high-recall semantic search^22^. This model maps both user queries and all text chunks in the knowledge base to a shared vector space, enabling efficient similarity computation. For each user query, the pipeline performs parallel searches across all curated databases (i.e., textbook, guidelines, and PubMed articles). Specifically, the system retrieves the top-5 most similar text chunks from each source using nearest-neighbor search in the ChromaDB vector database. Each retrieved chunk includes its associated metadata (e.g., source, chapter, and page number for textbooks, or article title and PMID for in PubMed), which is retained and passed to the downstream answer generation module. This design offers several key advantages. By searching each data source independently, the system achieves broad coverage of both literature and guidelines, significantly reducing the risk of missing relevant evidence. The inclusion of rich metadata with each retrieved chunk allows both the language model and human users to transparently trace the provenance of all supporting evidence, facilitating rigorous answer verification and auditability. Leveraging a high-dimensional embedding model, the system is better able to capture and discriminate the subtle nuances of complex medical knowledge, enabling more precise matching between user queries and relevant documents. Finally, dense embedding search enables rapid retrieval from large and heterogeneous knowledge bases, supporting efficient, near real-time performance even for demanding clinical scenarios.

### 3.4 Query Formulation and Inference Workflow

At the inference stage, our system is designed to maximize transparency, traceability, and user utility. After relevant chunks are retrieved from each data source, the language model (LLM) is instructed to process and structure the information in several specific ways. First, the LLM is required to explicitly indicate which retrieved passages it considers useful for answering the question, returning the corresponding metadata for each passage (e.g., source, chapter, section, and page number) so users can directly trace the evidence back to the original materials. For every cited passage, the model also assigns a confidence score from 1 to 10, reflecting both its trust in the passage’s accuracy and its judgment of how helpful the passage is in resolving the query. Including confidence scores serves several important purposes: passages with lower confidence can be automatically flagged, alerting users to possible ambiguities, conflicting evidence, or gaps in the knowledge base, while high-confidence citations offer users greater assurance that the answer is strongly supported. These scores also allow for later quantitative analyses, such as auditing the database for coverage and identifying areas in need of improvement. In addition to identifying useful sources, the model is prompted to synthesize the evidence and provide a natural-language rationale that details how the selected materials lead to the answer, or, if uncertainty remains, to explain the reasons for doubt. For multiple-choice settings, the model clearly states its selected option and provides reasoning for the rejection of other options. This workflow is designed to enhance transparency and trust by allowing users to verify both the final answer and the supporting evidence, and also to facilitate auditability for expert reviewers and support systematic data quality monitoring over time. Moreover, by surfacing both high- and low-confidence evidence, the system can help users recognize when answers are robust or when additional expert review may be warranted. All prompts and specific model instructions for this structured inference process are included in **Supplemental Table 1** to support reproducibility and facilitate comparisons with future methods.

### 3.5 Evaluation Protocol

To comprehensively evaluate our RAG pipeline’s ability to answer radiation oncology board-style questions, we employed the 2021 American College of Radiology (ACR) TXIT (The Radiation Oncology In-Training Examination) exam. The original dataset contained a total of 291 multiple-choice questions spanning key knowledge domains in radiation oncology. During data curation, we systematically removed any items requiring image interpretation or visual input, as our current RAG system is designed as text-only; after this filtering process, the final test set comprised 283 purely text-based questions. The ACR TXIT exam is a widely recognized standardized assessment administered annually to radiation oncology residents in the United States. Its content blueprint is designed by the American College of Radiology in alignment with the ABR (American Board of Radiology) Core Curriculum and covers a broad range e of essential topics – including radiation physics, radiobiology, anatomy, disease site management, treatment planning, and the latest evidence-based guidelines. TXIT is primarily used for benchmarking resident knowledge, guiding educational priorities for residency programs, and preparing trainees for the ABR board certification exam. Since its questions reflect current clinical standards and consensus guidelines, performance on TXIT is regarded as a strong proxy for a physician’s competence in contemporary radiation oncology. For each question in the dataset, we compared the answers generated by baseline large language models (OpenAI’s most advanced models without RAG augmentation) to those produced by our RAG-enhanced pipeline. Both models were prompted to select the best answer for each item, and responses were automatically compared to the official ACR answer key. Model accuracy was defined as the proportion of correctly answered questions relative to the oNicial answer key. We chose the TXIT exam as our primary evaluation instrument for several reasons. First, the diversity and breadth of topics ensures comprehensive assessment of the model’s generalizability and clinical knowledge. Second, the multiple-choice structure provides an objective and reproducible way to benchmark performance. Third, the real-world relevance and rigorous construction of TXIT questions allow our results to directly reflect the system’s potential value as a clinical decision support tool or an educational assistant in radiation oncology. Lastly, by excluding image-based questions, we focus specifically on the text-understanding and reasoning capabilities of our system, which are the core strengths of LLMs and RAG architectures.

### 3.6 Implementation Details

All data preprocessing, embedding generation, and retrieval operations were implemented in Python (version 3.12), using ChromaDB (version 1.0.12), the OpenAI API, and supporting libraries. The complete pipeline, from initial text fragment ingestion to automated answer evaluation, was managed via python scripts. Model evaluation covered several major versions of OpenAI’s large language models, including GPT-4 (released March 2023), GPT-4o (released May 2024), GPT-4.1 (released April 2025), and GPT-5 (released August 2025). For each model, both RAG-augmented and non-RAG baselines were tested under consistent settings. The typical processing time per question, from query formulation to final majority voting, was 5-10 seconds, depending on model size and retrieval scope. All results, including accuracy metrics and logs, were recorded automatically, ensuring the reproducibility of the experimental workflow. Further details and reproducible scripts are available upon request.

## 4 Results

### 4.1 Accuracy Improvement with Retrieval-Augmented Generation

**Figure 3** presents the comparative accuracy of four OpenAI large language models (GPT-4-turbo, GPT-4o, GPT-4.1, and GPT-5) across two answer generation strategies: direct response and retrieval-augmented generation (RAG). The results demonstrate two key findings regarding the impact of external knowledge integration. First, the magnitude of improvement from RAG is greatest for earlier-generation or lighter-weight models such as GPT-4-turbo, where accuracy increased by over 5 percentage points. This suggests that when a model’s parametric knowledge is less comprehensive or up-to-date, the ability to retrieve and condition on authoritative external data can dramatically mitigate knowledge gaps and significantly enhance performance. In contrast, more advanced models (e.g., GPT-4.1 and GPT-5) exhibit smaller absolute gains: GPT-4.1 accuracy improved from 85.2% to 87.3% and GPT-5 from 89.3% to 91.5%, indicating that as language models become more powerful and better pre-trained, the marginal benefit of retrieval diminishes but is not eliminated. This pattern reveals a complementary relationship: RAG especially benefits earlier or resource-constrained models, while still offering incremental improvements to state-of-the-art systems. An additional noteworthy finding is that the retrieval-augmented version of an older model (e.g., GPT-4-turbo with RAG, 79.38%) can outperform or match the direct performance of a much newer or larger model (e.g., GPT-4o without RAG, 77.3%). Similarly, GPT-4o with retrieval augmentation (82.47%) nearly reaches the level of GPT-4.1 without retrieval (85.2%), while GPT-4.1 with retrieval (87.3%) evinces similar performance to unaugmented GPT-5 (89.3%). This demonstrates the “leapfrogging” effect of retrieval augmentation: supplementing models with timely and targeted external information can close or even invert the performance gap between model generations. In effect, RAG serves as a force multiplier, enabling even smaller or older models to achieve results on par with newer, more computationally intensive models in specialized domains such as radiation oncology. Taken together, these findings highlight both the present and future significance of RAG in clinical AI applications. Not only does RAG enable current models to operate at a higher evidence-based standard, but it also provides a scalable pathway for keeping deployed models up-to-date, even as the underlying architectures and training data inevitably lag behind the latest clinical knowledge. This underscores the value of retrieval-augmented strategies for both robustness and long-term sustainability in medical AI deployment.

### 4.2 Ablation Study: Impact of Individual Knowledge Sources

Ablation analyses were conducted to isolate the contributions of individual knowledge sources in the retrieval-augmented generation (RAG) pipeline. As summarized in **Table 1**, accuracy and mean confidence were recorded while Gunderson’s textbook, NCCN Guidelines, and randomly sampled PubMed Central (PMC) articles were selectively enabled or disabled.

**Table 1:** Ablation study results comparing accuracy and mean confidence scores across different source combinations and LLM architectures.

| Knowledge Source |  |  | GPT-5 |  | GPT-4.1 |  | GPT-4o |  | GPT-4-turbo |  |
| --- | --- | --- | --- | --- | --- | --- | --- | --- | --- | --- |
| Textbook | NCCN | PMC | Acc (%) | Conf. | Acc (%) | Conf. | Acc (%) | Conf. | Acc (%) | Conf. |
| ✓ | ✓ | ✓ | 90.96 | **7.36** | 85.57 | 7.16 | 81.79 | **7.16** | **79.38** | **6.57** |
| ✓ | ✓ |  | **91.5** | 7.08 | **87.3** | **7.22** | **82.47** | 6.67 | 77.32 | 6.33 |
| ✓ |  | ✓ | 90.62 | 7.02 | 84.1 | 7.14 | 82.13 | 7.14 | 76.92 | 6.51 |
| ✓ |  |  | 90.78 | 7.18 | 85.91 | 7.12 | 81.91 | 6.53 | 77.91 | 6.23 |
|  | ✓ | ✓ | 90.41 | 7.12 | 84.2 | 6.10 | 82.13 | 6.13 | 76.73 | 6.20 |
|  | ✓ |  | 90.88 | 7.24 | 85.2 | 7.13 | 82.82 | 7.13 | 78.81 | 6.54 |
|  |  | ✓ | 90.07 | 6.05 | 84.57 | 6.47 | 78.4 | 4.79 | 75.24 | 5.32 |
|  |  |  | 89.3 | – | 85.2 | – | 77.3 | – | 73.1 | – |

**Figure 3:**
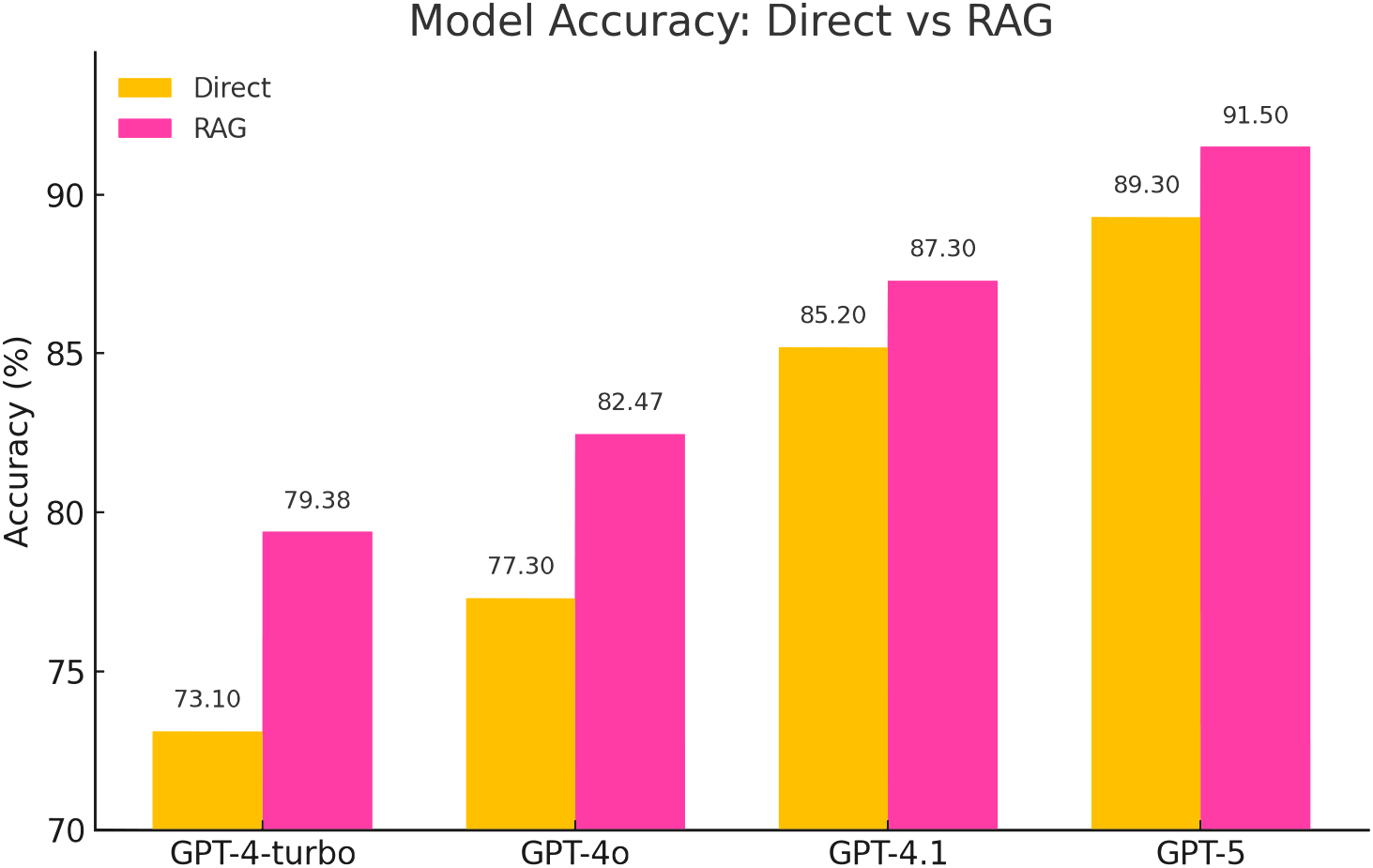
Model accuracy for direct answer versus retrieval-augmented generation (RAG, using textbook and guidelines). Incorporation of external knowledge consistently improves performance across all models.

Across all evaluated models (GPT-5, GPT-4.1, GPT-4o, and GPT-4-turbo), configurations including the textbook and NCCN were found to yield consistently higher accuracy and confidence. Under the textbook + NCCN setting, GPT-5 achieved the overall best performance of 91.5% with a mean confidence score of 7.08, representing a 2.2 percentage-point gain over its direct setting (89.3%). GPT-4.1 and GPT-4o reached 87.3% (7.22) and 82.47% (6.67), respectively, under the same two-source configuration.

When PMC articles were also included (textbook + NCCN + PMC), accuracy and confidence were generally attenuated for higher-capacity models: GPT-5 decreased from 91.5% (7.08) to 90.96% (7.36), and GPT-4.1 from 87.3% (7.22) to 85.57% (7.16). On the other hand, the lighter GPT-4-turbo model showed a modest improvement with the inclusion of PMC (accuracy increased from 77.32% to 79.38%, confidence from 6.33 to 6.57). Inclusion of PMC reduced performance of GPT-4o compared to textbook + NCCN (accuracy decreased from 82.47% to 81.79%); however, in contrast to other models, the GPT-4o textbook + NCCN model slightly outperformed the textbook alone (81.91% with textbook-only vs. 82.13% with textbook + PMC).

Due to the heterogeneous quality of information in PMC, adding it to the set of retrieval knowledge sources, in general, reduced performance. However, it is notable that all models except GPT-4.1 exhibited improved performance using PMC-only retrieval compared to direct runs without RAG.

Taken together, the results indicate that curated, consensus-based sources (textbook and NCCN) constitute the primary drivers of accuracy and reliable confidence, whereas indiscriminate inclusion of broad literature (PMC) can introduce ambiguity and reduce stability. Nevertheless, targeted primary literature can occasionally provide incremental benefits, particularly for earlier or lighter-weight models, while advanced models such as GPT-5 remain most reliable when anchored by curated sources.

### 4.3 Error Analysis via Confidence Scores

To evaluate the model’s ability to quantify the usefulness and reliability of retrieved documents within our RAG pipeline, confidence scores were analyzed for correct and incorrect responses across different GPT versions (**Figure 4**). The confidence score (1-10) reflects both the perceived helpfulness of retrieved evidence and the model’s certainty in the synthesized answer. As shown, GPT-5 exhibits the largest separation between confidence on correct versus incorrect answers (7.36 vs. 4.12), followed by GPT-4.1 (7.40 vs. 4.30), GPT-4o (7.12 vs. 5.10), and GPT-4-turbo (6.85 vs. 5.70). Similarly, when confidence score is treated as a threshold for classifying responses as “correct” or “incorrect” and receiver operating characteristic (ROC) curves plotted, GPT-5 confidence scores most accurately reflect the truth value of responses (AUC = 0.91), with the discriminatory performance declining for the less advanced models.

**Figure 4:**
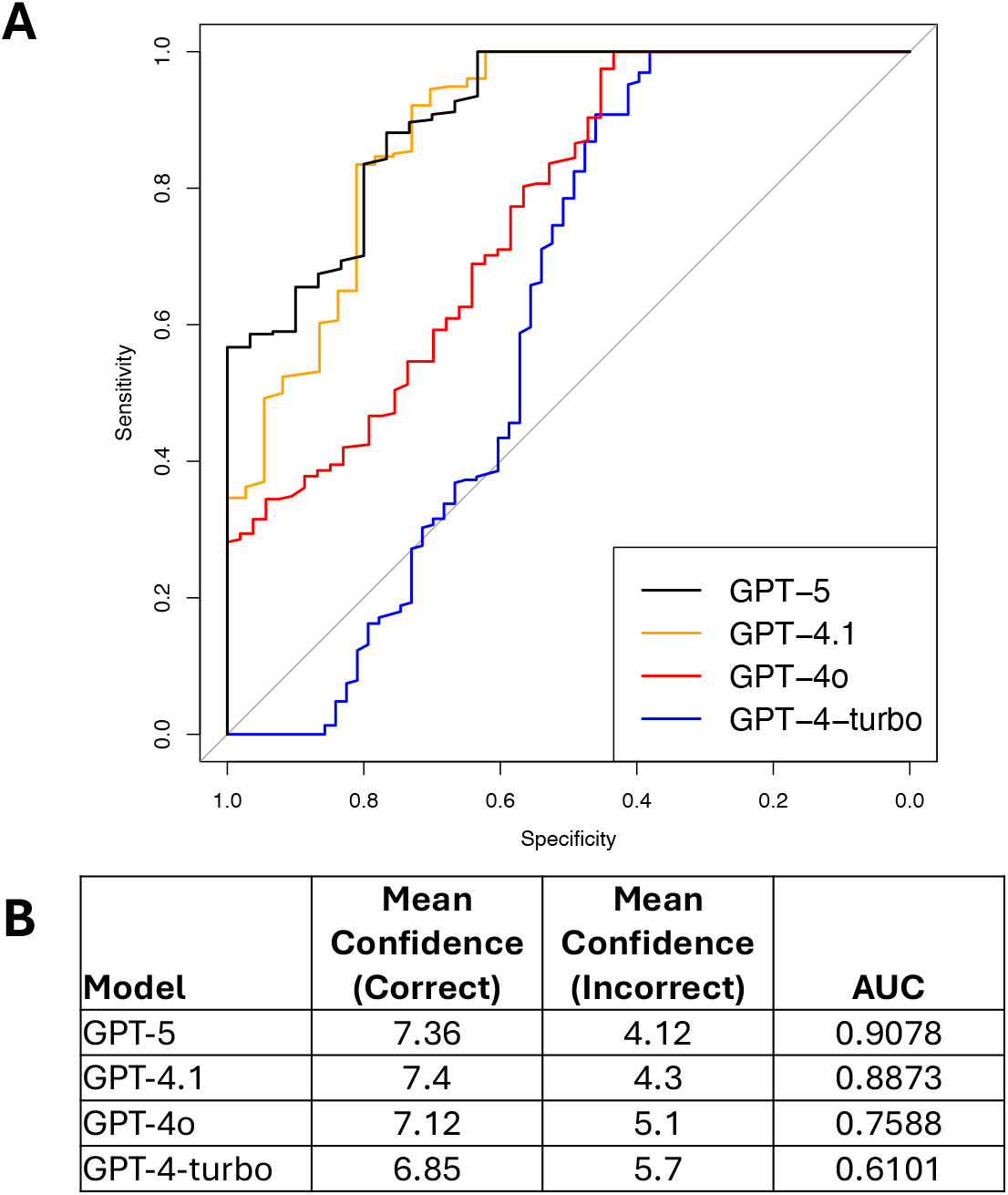
Ability of model-generated confidence scores to discriminate correct and incorrect responses across different GPT models. **(A)** ROC curves using varying confidence score thresholds to classify responses as “correct” or “incorrect” for the 4 different GPT models tested. **(B)** Table showing the mean confidence scores for correct and incorrect responses, as well as the area under the ROC curve (AUC) from panel A for each model.

From a clinical deployment perspective, the separation in scores between correct and incorrect answers is actionable. For example, a decision threshold around 5 would flag the average incorrect predictions of GPT-5 and GPT-4.1 (means < 5) for additional human verification, while many errors from GPT-4o and GPT-4-turbo (means > 5) might not be automatically flagged without a stricter policy. Overall, the trend indicates that more advanced models, when integrated with a structured RAG pipeline, not only improve accuracy but also better differentiate the sufficiency and reliability of supporting evidence through their confidence behavior.

## 5 Discussion

In this study, we have demonstrated that large language models augmented with our novel RAG-based workflow drawing on a well-curated, up-to-date expert knowledge base are able to answer questions in the highly specialized field of radiation oncology with a high level of accuracy. When benchmarking models’ ability to correctly answer purely text-based questions from the 2021 ACR TXIT examination (**Figure 3**), older, more lightweight models derived the greatest benefit from RAG-augmentation (GPT-4-turbo accuracy increased from 73.1% to 79.4%), but even the latest GPT-5 model showed substantial improvement, increasing its accuracy from 89.3% to a superlative 91.5%. This general applicability is encouraging, and the strong benefit derived by lighter weight models is of particularly relevance in the healthcare and clinical support setting, where data privacy concerns tend to favor reliance on smaller, local models.

A notable strength of our approach in developing practical models for clinical support and medical education is the provision of confidence scores, by means of which our multi-agent RAG pipeline reports to users an assessment of its ability to correctly answer questions based on the available data. This allows users to readily identify when a query has extended beyond the expert knowledge base provided to a model or may touch on a controversial point where sources differ. In addition, each answer is tied directly to a set of auditable, confirmable citations that allow users to directly trace each answer back to a verified source.

The representative questions in **Table 2** demonstrate the strengths and potential pitfalls of our retrieval-augmented generation (RAG) pipeline in answering radiation oncology board-style questions.

**Table 2:** Comparative analysis of success and failure cases for the RAG pipeline. In the Success Case, the RAG-augmented model was able to correctly answer a question with high confidence that was missed by the direct model by basing its answer on relevant text retrieved from an up-to-date NCCN guideline. In the Failure Case, the RAG model retrieved no text relevant to the question; with low confidence in the text, the model relied on embedded knowledge, returning the same incorrect answer as the direct model.

| Aspect | Success Case | Failure Case |
| --- | --- | --- |
| Question | 4-year local control rate after SBRT for primary renal cell carcinoma | Top priority in maintaining trial integrity using historical controls in Phase III trials |
| Direct Answer | Incorrect (81-90%) | Incorrect (Control type I error) |
| RAG Answer | Correct (91-100%) | Incorrect (Control type I error) |
| Confidence Score | 9.0 (High confidence) | 4.0 (Low confidence) |
| Provided Explanation | Multiple studies, including a pooled analysis by the International Radiosurgery Oncology Consortium for Kidney (IROCK), report 4-year local control rates of approximately 97-98% after SBRT for primary renal cell carcinoma. This places the correct answer in the highest range provided. | When using historical controls in a Phase III trial, the top priority is to control the type I error. This is because historical controls can introduce biases due to differences in patient populations, treatments, and data collection over time. Without strict control of type I error, the risk of falsely concluding a treatment effect (false positive) increases, threatening trial validity and integrity. The other options (A, B, C) are not primary considerations for maintaining trial integrity in this context. |
| Retrieved Text | Precise NCCN guideline detailing exact figures from pooled analyses | Irrelevant information regarding Phase II trials, lacking direct Phase III trial relevance |

In the success case, the RAG pipeline successfully leveraged a specific NCCN guideline, significantly improving the response by providing an accurate, high-confidence answer (confidence score: 9.0). This case demonstrates the core strength of our RAG pipeline: the retrieval of exact and authoritative information directly relevant to the clinical question, improving both precision and interpretability. In contrast, in the failure case, the retrieved supporting information was not directly relevant to the specific query regarding Phase III trials, instead offering details only pertinent to Phase I and II trials. This irrelevant retrieval was tagged with a low confidence score (4.0), signaling the potential unreliability of the provided answer and causing the model to rely on its embedded knowledge to produce an answer, resulting in the same incorrect response as the direct model.

The contrast between these cases underscores the importance of the quality and relevance of the retrieved information. Although high-quality and directly relevant guidelines significantly boost performance and confidence, irrelevant or indirectly related information introduces ambiguity and can mislead the inference model. This point was directly demonstrated in our tests incorporating 1,000 radiation oncology papers randomly selected from PubMed into the expert knowledge base for our RAG models: contrary to the conventional wisdom that, when training language models, “more is better”, the introduction of a large quantity of uncurated, although nominally relevant, text into the knowledge base reduced model performance (**Table 1**).

In practical use cases with a well-curated knowledge base, confidence scores serve as crucial indicators of answer reliability. Physicians and researchers can therefore use low confidence scores as an early warning system, prompting careful manual review or supplementary consultation. Low confidence cases further emphasize the importance of ongoing refinement of the knowledge base, highlighting the need for high precision in the selection of resource materials, particularly in domains where nuanced distinctions between trial phases or specific clinical conditions are critical.

Making use of the auditing power provided by our RAG pipeline, we manually reviewed the cited material relied on by models when returning incorrect responses in order to understand the causes of errors. *To better characterize and systematically address these failure modes, we classified observed errors into four major types:*

### 1. Knowledge Absence

The knowledge base does not contain the required information.

### 2. Retrieval Failure

The knowledge exists in the database but is not retrieved.

### 3. Utilization Failure

The correct information is retrieved but not used appropriately in the response.

### 4. Controversial/Ambiguous Question

The question lacks a clear, evidence-based answer or is subject to ongoing clinical debate.

See **Supplemental Table 2** for a detailed breakdown of representative failure cases across these error types.

One notable shortcoming of this and similar studies is that, to facilitate evaluation of model performance in answering expert queries on topics related to radiation oncology, we relied on a multiple-choice test (2021 ACR TXIT). While this structure makes the unambiguous identification of correct and incorrect answers trivial, it is removed from the way such a model would be used in actual clinical support or medical education scenarios, in which open-ended questions would be posed to the model. Our follow-on effort to this work will assess RAG-augmented model performance on such open-ended questions, including in simulated clinical scenarios, with results evaluated by a panel of domain experts.

## 6 Conclusion and Future Work

In this study, we demonstrated that retrieval-augmented generation (RAG) pipelines substantially improve both the factual accuracy and citation reliability of large language models (LLMs) when answering complex, board-style radiation oncology questions. By integrating authoritative reference materials, such as clinical guidelines and textbooks, into the answer generation process, our multi-agent RAG system not only outperformed direct LLM baselines across all model variants, but also provided transparent evidence trails for human verification. We further highlighted that careful construction, curation, and ongoing maintenance of the knowledge base are essential to achieving optimal system performance, as both the accuracy and reliability of answers depend critically on the quality and relevance of retrieved information. Moreover, the use of confidence scores and supporting evidence offers practical tools for clinicians to assess the trustworthiness of automated recommendations, thereby promoting safer and more interpretable deployment in real-world clinical settings.

Looking ahead, several directions can further enhance RAG-based medical AI systems. First, the deployment of local, open-source language models may address data privacy concerns and improve accessibility for institutions with limited access to commercial APIs. Second, expanding the agent-based architecture to incorporate more sophisticated reasoning mechanisms – such as explicit chain-of-thought (CoT) or tool-augmented inference – could further boost answer quality, especially for complex multi-step questions. Finally, developing user-facing applications that seamlessly integrate into clinical workflows and electronic health records will be critical for real-world adoption. Such tools should prioritize user experience, auditability, and regulatory compliance, ensuring that clinicians can interact intuitively with AI systems while retaining full control over the final clinical decision-making process.

## Data Availability

All data produced in the present study are available upon reasonable request to the authors

