## Supplemental Data for "Development of a RAG-based Expert LLM for Clinical Support in Radiation Oncology"

| Prompt Type | Prompt Content |
|---|---|
| A. RAG Inference | You are an expert in Radiation Oncology. The user asked:<br>Question: “{user question}”<br>Below is some text retrieved from external documents (which might be incomplete or irrelevant):<br>{retrieved context}<br>Instructions:<br>1. Provide a concise, medically accurate answer based on your expert knowledge of Radiation Oncology.<br>2. If the retrieved text is relevant and correct, incorporate it into your reasoning.<br>3. If the retrieved text seems contradictory, incomplete, or irrelevant, rely primarily on your own expertise.<br>4. If there is insufficient data, provide your best answer along with any disclaimers.<br>5. Ensure your final answer in ABCD only contains “A”, “B”, “C”, “D” and no other characters.<br>6. Provide (1-10) as a score for how helpful the retrieved document was in your answer.<br>7. Your explanation should be short and concise.<br>8. Only cite passages that are directly helpful for answering the question. Ignore irrelevant content.<br>9. For each passage you cite, output its source metadata and how helpful it was (on a scale of 1 to 10). |
| B. Baseline (Direct LLM) | You are an expert in Radiation Oncology. Please provide a concise, medically accurate answer to the following question.<br>Question: “{user question}”<br>Instructions:<br>1. Provide your final answer in ABCD format (only the letter: A, B, C, or D).<br>2. Give a brief explanation, ensuring it is short and to the point. |

**Supplemental Table 1: Prompt Templates.** This table documents the exact prompt templates employed in the RAG inference stage and for baseline (direct LLM) model evaluation. These prompts were provided to the large language model to ensure consistent and reproducible outputs across experiments. Metadata placeholders (e.g., {user question} and {retrieved context}) were automatically filled by the pipeline at runtime.

| Question | Error Type | Detailed Analysis |
| --- | --- | --- |
| When using historical controls in a Phase III trial, what is the top priority to maintain trial integrity? | Retrieval Failure | **Question:** Trial integrity priorities (options: sample size, duration, remove placebo, control type I error).<br>**RAG Answer:** Control type I error (incorrect, correct was remove placebo).<br>**Context:** RAG retrieved general text on trial design but not specific to trial integrity with historical controls.<br>Commentary: The relevant guideline on historical controls was not surfaced, resulting in the wrong focus. Improved retrieval could have avoided this error. |
| Which patient would gain the greatest benefit in local control from RT (in DCIS scenarios)? | Utilization Failure | **Question:** Four DCIS scenarios.<br>**RAG Answer:** Larger, intermediate grade DCIS post-lumpectomy (B, incorrect; correct was high-grade, small, post-lumpectomy [A]).<br>**Context:** RAG retrieved multiple trials on RT for DCIS, but model failed to correctly synthesize and apply the context to prioritize the right risk features. |
| Which clinical finding is characteristic of locally advanced breast cancer? | Utilization Failure | **Question:** Four findings, select for LABC.<br>**RAG Answer:** Nipple retraction (C, incorrect; correct: palpable supraclavicular node [D]).<br>**Context:** Text included many LABC features, but model failed to distinguish between local invasion and nodal spread. |
| What is the TMN classification in a patient that presents with a 5 cm, grade 3 breast cancer with matted axillary lymph nodes, received neoadjuvant chemotherapy, and had a pCR at time of MRM? | Utilization Failure | **Question:** Staging before and after neoadjuvant therapy.<br>**RAG Answer:** cT3N2aM0, ypT0N0Mx (D, incorrect; correct: cT2N2aM0, ypT0N0).<br>**Context:** Retrieved general staging info, but failed to fully apply pathologic response notation or tumor size criteria. |
| What is the BEST treatment for a 55 year-old female who underwent breast-conserving surgery for a pT1cN1mi cM0 ER+ HER2- breast cancer and 21 gene recurrence score of 22? | Controversial Question | **Question:** Best adjuvant therapy given clinical/pathologic factors and genomic risk.<br>**RAG Answer:** Chemotherapy + RT + endocrine therapy (C, incorrect; correct: RT + endocrine only).<br>**Context:** NCCN and recent RCTs provide divided recommendations for this patient profile. Model reflected ambiguity. |

| | | |
|---|---|---|
| Which laboratory test abnormality is MOST frequently associated with a rectal adenocarcinoma? | Knowledge Absence | **Question:** Pick one of CEA, CA19-9, CA125, AFP.<br>**RAG Answer:** CEA (A, incorrect; correct: CA125 [C]).<br>**Context:** Retrieved context discussed CEA for colorectal, but did not specify for rectal cancer. The knowledge base lacked a clear statement for rectal adenocarcinoma. |
| Which of the following best defines non-classic radiation-induced liver disease (RILD)? | Retrieval Failure | **Question:** Diagnostic criteria for non-classic RILD.<br>**RAG Answer:** Increase in Child-Pugh score by 2+ (B, incorrect; correct: ALBI grade up by 1+).<br>**Context:** RAG found general RILD descriptions but did not surface the specific ALBI definition, despite database coverage. |
| Which of the following would be the LEAST acceptable fractionation regimen for definitive RT for a 4 cm hepatocellular carcinoma? | Utilization Failure | **Question:** Four dose/fraction regimens.<br>**RAG Answer:** Picked 60 Gy/3 fx as least acceptable (A, incorrect; correct: 40 Gy/5 fx).<br>**Context:** RAG retrieved data on SBRT regimens, but failed to apply BED and clinical acceptability for HCC. |
| What is the preferred first-line systemic therapy for unresectable, advanced hepatocellular carcinoma? | Knowledge Absence | **Question:** Options included atezolizumab/bevacizumab, lenvatinib, sorafenib, regorafenib.<br>**RAG Answer:** Sorafenib (incorrect; correct: atezolizumab/bevacizumab per most recent guidelines).<br>**Context:** RAG missed most recent updates to NCCN/ASCO, likely because knowledge base had not yet incorporated the newest guideline. |
| Which chemotherapy regimen is preferred for relapsed glioblastoma? | Retrieval Failure | **Question:** Multiple options, requires current NCCN.<br>**RAG Answer:** Temozolomide rechallenge (incorrect; correct: lomustine-based regimen per guideline).<br>**Context:** Knowledge base contained relevant guideline, but retrieval surfaced older or less relevant context. |

**Supplemental Table 2: Detailed Failure Cases and Error Taxonomy.** This table presents representative failure cases for the RAG pipeline, each mapped to one of four major error types: (1) Knowledge Absence, (2) Retrieval Failure, (3) Utilization Failure, and (4) Controversial/Ambiguous Question. For each, we include the clinical question, answer choices, the model's answer, correct answer, retrieved context (summarized), and analytic commentary.
